# Continuous Associations Between Remote Self-Administered Cognitive Measures and Imaging Biomarkers of Alzheimer’s Disease

**DOI:** 10.1101/2024.03.01.24303616

**Authors:** Elizabeth A. Boots, Ryan D. Frank, Winnie Z. Fan, Teresa J. Christianson, Walter K. Kremers, John L. Stricker, Mary M. Machulda, Julie A. Fields, Jason Hassenstab, Jonathan Graff-Radford, Prashanthi Vemuri, Clifford R. Jack, David S. Knopman, Ronald C. Petersen, Nikki H. Stricker

**Author notes:** Corresponding Author: Nikki H. Stricker, Ph.D., Mayo Clinic, 200 First Street SW, Rochester, MN 55905, (email). Copyright 2024 Mayo Foundation for Medical Education and Research. A poster presentation of early/partial data from this work in a smaller sample size was presented at the Alzheimer’s Association International Conference (July 2023). Another portion of this work was submitted as an abstract for the American Psychological Association Annual Convention (August 2024).

## Abstract

**Background:** Easily accessible and self-administered cognitive assessments that can aid early detection for Alzheimer’s disease (AD) dementia risk are critical for timely intervention.

**Objectives/Design:** This cross-sectional study investigated continuous associations between Mayo Test Drive (MTD) – a remote, self-administered, multi-device compatible, web-based cognitive assessment – and AD-related imaging biomarkers.

**Participants/Setting:** 684 adults from the Mayo Clinic Study of Aging and Mayo Clinic Alzheimer’s Disease Research Center participated (age=70.4±11.2, 49.7% female). Participants were predominantly cognitively unimpaired (CU; 94.0%).

**Measurements:** Participants completed (1) brain amyloid and tau PET scans and MRI scans for hippocampal volume (HV) and white matter hyperintensities (WMH); (2) MTD remotely, consisting of the Stricker Learning Span and Symbols Test which combine into an MTD composite; and (3) in-person neuropsychological assessment including measures to obtain Mayo Preclinical Alzheimer’s disease Cognitive Composite (Mayo-PACC) and Global-z. Multiple regressions adjusted for age, sex, and education queried associations between imaging biomarkers and scores from remote and in-person cognitive measures.

**Results:** Lower performances on MTD were associated with greater amyloid, entorhinal tau, and global tau PET burden, lower HV, and higher WMH. Mayo-PACC and Global-z were associated with all imaging biomarkers except global tau PET burden. MCI/Dementia participants showed lower performance on all MTD measures compared to CU with large effect sizes (Hedge’s g’s=1.65-2.02), with similar findings for CU versus MCI only (Hedge’s g’s=1.46-1.83).

**Conclusions:** MTD is associated with continuous measures of AD-related imaging biomarkers, demonstrating ability to detect subtle cognitive change using a brief, remote assessment in predominantly CU individuals and criterion validity for MTD.

## Introduction

Novel treatments targeting the underlying pathophysiology of Alzheimer’s disease (AD) are now available,^1^ with ongoing and future clinical trials likely to result in even more treatment options for individuals with biomarker-confirmed AD.^2^ Given these new treatment options, along with existing behavioral approaches to prevent or slow the progression of the AD clinical syndrome,^3^ the need for easily accessible and widespread cognitive screening of older adults is more imperative than ever before as it allows for timely, early detection of cognitive change during a critical time window for treatment intervention. In particular, brief cognitive assessment tools that can be deployed remotely and self-administered have high utility for decentralized clinical trials of AD as well as for use in clinical settings, where these assessments can aid in triage to specialty clinics, inform treatment initiation, support inclusion, and inform cognitive progression in clinical trials.^4,5^ Development of brief, digital remote cognitive assessments that detect subtle cognitive change and are sensitive to AD-related pathological change also allow for greater accessibility compared with in-person neuropsychological testing and could allow for future comparison with plasma AD biomarkers, which are themselves poised to become highly accessible when compared with imaging and cerebrospinal fluid biomarkers.^6^

Preclinical Alzheimer’s Cognitive Composites (PACCs) are commonly utilized outcome measures in longitudinal studies and AD clinical trials and have prioritized inclusion of in-person cognitive measures sensitive to subtle cognitive changes observed in preclinical AD.^7^ To date, few remote digital assessments have been assessed as measures sensitive to subtle cognitive changes observed in preclinical AD in a comparable fashion to PACCs. One computerized cognitive assessment, the Computerized Cognitive Composite (C3), showed moderate correlations with the Alzheimer Disease Cooperative Study (ADCS)-PACC (r=0.39) and showed similar small cross-sectional group differences across amyloid positive (A+) and amyloid negative (A-) groups compared to the ADCS-PACC.^8^ However, although capable of being administered remotely, the C3 was completed in person with an administrator present. Additionally, there is evidence to suggest that PACCs can still be further optimized, with one study demonstrating that person-administered measures including processing speed and learning and memory were more sensitive to longitudinal cognitive decline than the PACC5 in individuals with emerging amyloid beta, prior to typical amyloid-positive thresholds.^9^ Thus, a remote, digital cognitive assessment that employs measures most sensitive to the earliest changes in both cognition and biomarkers of AD could provide a more scalable and accessible alternative that is comparable to current PACCs.

Mayo Test Development through Rapid Iteration, Validation, and Expansion (Mayo Test Drive, MTD) is a remote, self-administered, multi-device compatible, web-based cognitive assessment. ^10^ MTD provides more in-depth assessment of key cognitive domains that are sensitive to subtle cognitive change relative to typical cognitive screening measures and is specifically designed for remote assessment. MTD consists of the Stricker Learning Span (SLS), a novel computer adaptive verbal word list memory test,^10,11^ as well as the Symbols Test, an open-source measure of processing speed that requires visuospatial discrimination and correlates with measures of processing speed and executive functioning.^10^ MTD has certain advantages over PACCs in that it can be easily completed in a remote and self-administered fashion; we have previously shown that MTD has high usability in cohorts of older adults and can typically be completed in about 15 minutes.^10,12^ Prior work from our group has also demonstrated that the SLS is able to differentiate between AD biomarker-defined groups comparably to the in-person administered Auditory Verbal Learning Test (AVLT), including distinguishing participants who were A+ versus A-as well as those who were amyloid and tau positive (A+T+) versus amyloid and tau negative (A-T-).^11^ Notably, this work was done in a predominantly cognitively unimpaired (CU) sample, suggesting that the SLS can detect subtle objective cognitive decline that aids in differentiation of biomarker positive versus negative participants with no other overt clinical symptoms.

Given that combining measures of learning/memory and processing speed/executive function provides richer information regarding cognitive performance than either in isolation, it is important to understand how performance on a composite measure of MTD is related to biomarkers of AD in a continuous fashion, rather than by splitting into positive and negative groups. Demonstrating that MTD performance is associated with continuous measures of amyloid, tau, neurodegeneration, and vascular injury is in line with the Alzheimer’s Association research framework for biological diagnosis of AD^13^ (currently under revision), as well.

In addition to possible utility for detecting subtle cognitive change associated with AD biomarkers in a remote fashion, another potential advantage of MTD compared with PACCs is its ability to produce a raw composite score to facilitate clinical utility and generalizability across different samples. Currently, PACCs rely on z-scores that are study specific, where z-scores are typically referenced to the sample presented within the study, making interpretation of performance across studies more challenging.^14–16^ By contrast, in the current study we introduce an MTD raw composite score that can allow MTD to function like a Mini Mental Status Exam (MMSE) or Montreal Cognitive Assessment (MoCA). A raw score composite will facilitate development of easily understood and generalizable cut-offs that do not adjust away the effects of age, which is highly confounded with increased prevalence of AD biomarkers and other undetected neuropathologies^17,18^ and thereby decreases sensitivity to mild cognitive impairment (MCI).^19^ This MTD raw composite thus retains the benefits of a total raw score for screening purposes and can also be used in conjunction with normative data.^20,21^

The primary aim of this study was to investigate continuous associations between MTD and imaging biomarkers to examine the criterion validity of MTD and its subtests as a measure of AD-related cognitive change in a population-based study of aging. Specifically, we hypothesized that worse performances on MTD would be associated with greater amyloid and tau PET burden, lower hippocampal volumes (HV), and higher white matter hyperintensity (WMH) volume. A secondary aim of this study was to validate a new MTD raw composite variable that can be used independently of specific study samples. We hypothesized that the MTD raw composite variable would demonstrate a similar pattern of results compared to both an alternative MTD composite that uses a traditional z-score approach and to existing in-person cognitive composite measures in association with imaging biomarkers. Finally, we also report group difference effect sizes for MTD composite and subtest variables across diagnostic groups (CU, MCI, and dementia) to inform potential clinical utility and validity, as well as correlations between MTD and imaging biomarkers across diagnostic groups.

## Methods

### Participants

The majority of participants (98.5%) were from the Mayo Clinic Study of Aging (MCSA). The MCSA is a longitudinal, population-based study of aging for residents of Olmsted County, Minnesota. Participants are recruited via random sampling of age– and sex-stratified groups using the Rochester Epidemiology Project medical records-linkage system.^22^ Study visits, which occur every 15 months, involve a physician examination, study coordinator interview that includes the Clinical Dementia Rating (CDR^®^) instrument,^23^ and neuropsychological testing.^24^ After each study visit, a diagnosis of CU, MCI,^25^ or dementia^26^ is established after the examining physician, interviewing study coordinator, and neuropsychologist make independent diagnostic determinations and come to consensus agreement.^24^ All participants are also invited to undergo and sign informed consent for neuroimaging. Participants with MCI or dementia complete study visits every 15 months, whereas CU individuals complete visits every 30 months if age 50 or older, or every 60 months if younger than 50 years of age. The remainder of participants was recruited from the Mayo Clinic Alzheimer’s Disease Research Center (ADRC; Rochester, MN). This study was approved by the Mayo Clinic and Olmsted Medical Center Institutional Review Boards and conducted in accordance with the Declaration of Helsinki. Written consent was obtained from all participants for participation in the overall study protocols (MCSA or ADRC) and oral consent was obtained for participation in the ancillary study protocol by which MTD was obtained via remote data collection.

Initial inclusion criteria for this specific study consisted of completion of MTD and available neuroimaging data (MRI and/or PET) completed within 36 months of completion of MTD. Participants were included across all diagnostic categories (i.e., CU, MCI, and dementia). Most participants had both MRI and PET neuroimaging data available as well as complete in-person neuropsychological data; however, specific sample sizes varied across outcome measures (see *Table 1*).

**Table 1.** Participant Characteristics for the Full Sample as well as for Cognitively Unimpaired and Mild Cognitive Impairment/Dementia Participants.

| Characteristic* | Full Sample<br>N=684<sup>1</sup> | CU<br>N=643 | MCI/Dementia<br>N=41 | p<sup>†</sup> | Hedge's g |
| --- | --- | --- | --- | --- | --- |
| **Demographics** |  |  |  |  |  |
| Age at MCSA/ADRC visit, years | 70.363 (11.204) | 69.890 (11.111) | 77.786 (10.097) | <0.001 | 0.71 (0.40, 1.03) |
| Sex, N (%) Female | 340 (49.7%) | 318 (49.5%) | 22 (53.7%) | 0.60<sup>‡</sup> | -- |
| Education, years | 15.646 (2.381) | 15.719 (2.335) | 14.512 (2.812) | 0.002 | -0.51 (-0.83, -0.19) |
| Race, N (%) White<sup>2</sup> | 664 (97.1%) | 624 (97.0%) | 40 (97.6%) | 1.00<sup>§</sup> | -- |
| Ethnicity, N (%) Non-Hispanic<sup>3</sup> | 680 (99.4%) | 639 (99.4%) | 41 (100.0%) | 1.00<sup>§</sup> | -- |
| In-person visit to MTD, months | 0.737 (1.965) | 0.709 (1.896) | 1.169 (2.831) | 0.15 | 0.23 (-0.08, 0.55) |
| Imaging to MTD, months | 5.746 (11.393) | 6.113 (11.622) | -0.016 (3.492)<sup>4</sup> | <0.001 | -0.54 (-0.86, -0.23) |
| MTD completed in clinic, N (%) | 3 (0.4%) | 3 (0.5%) | 0 (0.0%) | 1.00<sup>§</sup> | -- |
| **Cognition: Mayo Test Drive<sup>5,7</sup>** |  |  |  |  |  |
| MTD-SBCr, n=680 | 105.221 (22.675) | 107.653 (20.171) | 66.314 (25.200) | <0.001 | -2.02 (-2.35, -1.68) |
| MTD-SBCz, n=680 | -0.072 (0.929) | 0.030 (0.815) | -1.718 (1.092) | <0.001 | -2.10 (-2.44, -1.76) |
| SLS Sum of Trials, n=681 | 74.023 (18.747) | 75.833 (17.174) | 45.025 (19.265) | <0.001 | -1.78 (-2.11, -1.45) |
| SYM, n=681 | 3.472 (1.313) | 3.337 (1.067) | 5.623 (2.513) | <0.001 | 1.91 (1.57, 2.24) |
| SYM<sub>aw</sub>, n=681 | 31.219 (6.849) | 31.839 (6.065) | 21.289 (10.338) | <0.001 | -1.65 (-1.98, -1.32) |
| **Cognition: In-Person Measures<sup>6,7</sup>** |  |  |  |  |  |
| Mayo-PACC, n=661 | -0.065 (0.844) | 0.026 (0.738) | -1.688 (0.978) | <0.001 | -2.28 (-2.64, -1.92) |
| Global Cognition z, n=629 | 1.419 (1.004) | 1.518 (0.887) | -0.871 (0.819) | <0.001 | -2.70 (-3.12, -2.28) |
| AVLT Sum of Trials, n=668 | 65.177 (18.759) | 66.979 (17.568) | 35.289 (11.130) | <0.001 | -1.83 (-2.18, -1.49) |
| Trail Making Test B, n=669 | 77.166 (43.657) | 72.472 (34.875) | 157.351 (83.977) | <0.001 | 2.17 (1.82, 2.52) |
| Digit Symbol Coding, n=658 | 51.299 (12.650) | 52.060 (12.222) | 34.179 (9.843) | <0.001 | -1.47 (-1.86, -1.09) |
| STMS, n=677 | 35.581 (2.624) | 35.962 (1.905) | 29.500 (4.449) | <0.001 | -3.02 (-3.38, -2.67) |
| **Neuroimaging Metrics<sup>5</sup>** |  |  |  |  |  |
| Amyloid PET Meta-ROI SUVR, n=670 | 1.549 (0.375) | 1.527 (0.341) | 1.890 (0.637) | <0.001 | 0.99 (0.67, 1.32) |
| Tau PET Meta-ROI SUVR, n=667 | 1.209 (0.140) | 1.197 (0.098) | 1.389 (0.374) | <0.001 | 1.45 (1.13, 1.78) |
| Tau PET EC-ROI SUVR, n=667 | 1.134 (0.159) | 1.120 (0.130) | 1.360 (0.322) | <0.001 | 1.62 (1.29, 1.95) |
| Hippocampal Volume z-score, n=680 | -0.343 (0.686) | -0.283 (0.611) | -1.292 (1.023) | <0.001 | -1.57 (-1.90, -1.24) |
| % WMH Volume, ln, n=665 | -0.645 (0.904) | -0.680 (0.884) | -0.111 (1.047) | <0.001 | 0.64 (0.32, 0.95) |

\*Values are presented as Mean (Standard Deviation) unless otherwise noted.
† All between-group comparison are T-tests unless otherwise indicated.
‡ ☐ Chi-square ☐ ☐ ☐
§ ☐ Fisher
<sup>1</sup> n=674 Mayo Clinic Study of Aging (MCSA); n=10 Mayo Alzheimer's Disease Research Center (ADRC) Rochester, MN.
<sup>2</sup> n=7 Asian, n=5 Black, n=8 Missing/Unknown
<sup>3</sup> n=2 Missing/Unknown
<sup>4</sup> Negative value indicates this group on average had imaging visits prior to MTD completion. By study design, individuals with MCI/dementia complete imaging every 15 months, whereas CU individuals complete imaging every 30 months if age 50 or older, or every 60 months if younger than 50 years of age.
<sup>5</sup> Mayo Test Drive and Neuroimaging are independent of diagnosis (data not considered for consensus diagnosis).
<sup>6</sup> Results of in-person cognitive measures are considered for consensus diagnosis.
<sup>7</sup> Results remained significant when adjusting for age, sex and education (all $p$ 's $\leq .003$ ).
Note: ADRC = Alzheimer's Disease Research Center; AVLT = Auditory Verbal Learning Test; CU = Cognitively Unimpaired; EC = entorhinal cortex; Mayo-PACC = Mayo Preclinical Alzheimer's Disease Cognitive Composite; MCI = Mild Cognitive Impairment; MCSA=Mayo Clinic Study of Aging; MRI = Magnetic Resonance Imaging; MTD = Mayo Test Drive; MTD-SBCr = Mayo Test Drive Screening Battery Composite raw; MTD-SBCz = Mayo Test Drive Screening Battery Composite z; p = p-value; PET = Positron Emission Tomography; ROI = Region of Interest; SLS = Stricker Learning Span; STMS = Short Test of Mental Status; SUVR = Standard Uptake Volume Ratio; SYM = Symbols Test average correct item response time; SYM<sub>aw</sub> = Symbols Test accuracy-weighted average correct item response time; WMH = White Matter Hyperintensities.

### Self-Administered, Remote Cognitive Measures

All participants completed MTD^10,11^ approximately a few weeks following their in-person MCSA or ADRC visit (0.74 months on average). Participants were invited via email to complete the measures remotely, and nearly all participants completed MTD remotely and unsupervised. Three participants completed MTD in clinic per their request (0.4% of study sample). MTD is comprised of the SLS and the Symbols Test, from which composite measures are calculated, as described below.

### Stricker Learning Span (SLS)

The SLS is a computer adaptive list learning task; please see prior publications for a detailed review of test procedures.^10,11,27^ Briefly, single words are visually presented sequentially across five learning trials. After each list presentation, memory for each word on the word list is assessed with four-choice recognition. The SLS begins with eight items and then the number of words either stays the same, increases, or decreases depending on performance across each trial (range 2-23 words by trial five). The delay trial of the SLS occurs following completion of the Symbols Test and presents all items that were previously presented on any SLS learning trial. The SLS Sum of Trials, the primary outcome variable from this task, is calculated as the sum of words correctly remembered from Trials 1-5 and the Short Delay trial. Additional secondary SLS variables include SLS Maximum Learning Span, SLS Trials 1-5 Total, and SLS Short Delay Total.

### Symbols Test

The Symbols Test is an open-source measure of processing speed with previously demonstrated validity and reliability.^28,29^ Processing speed measures are routinely incorporated into composite cognitive measures designed to be sensitive to early preclinical changes due to their known sensitivity to cognitive aging, AD, and other neurodegenerative disorders.^10,30^ For each item, participants identify which of two symbol pairs on the bottom of a screen matches one of three symbol pairs presented at the top of the screen. The original version is part of the Ambulatory Research and Cognition (ARC) app^31^ and includes up to 28 12-item trials taken over the course of seven consecutive days. The number of trials for this shortened version (four sequential trials) was selected based on data showing that between-person reliability passes a 0.80 reliability threshold after 2 trials.^29^ The primary outcome variable used in other studies is average correct item response time (SYM, sec) across four 12-item trials. Because one goal of MTD is clinical use, we include additional variables. First, although the primary outcome variable only includes correct items, we have observed that response times tend to be consistent, whether accurate or inaccurate. Clinically, there may also be some utility in understanding how accuracy influences performance. For this reason, we created a modification of the SYM variable that additionally accounts for accuracy, SYM_aw_, which is defined below.

### MTD Composite Measures

We included two MTD composite variables in this study. The first, MTD screening battery composite z (MTD-SBCz), is a composite based on a traditional z-score approach that has been previously demonstrated to show good diagnostic accuracy for MCI in preliminary data.^32^ This composite is the average of the z-scores of the following four variables, which were created from all participants available at the time of the data freeze for this analysis, including those without imaging (N=1447; 95.5% CU): SLS Maximum Learning Span, SLS Trial 1-5 Total, SLS Short Delay Total, and SYM (reversed such that higher scores reflect better performance).

The second MTD composite variable is the MTD screening battery composite raw (MTD-SBCr). For this composite, we aimed to create a composite on a raw score scale that performed similarly to the MTD-SBCz but also factored in accuracy on the Symbols test, in addition to speed, to help capture variance observed in participants with low accuracy. Given that SYM accuracy has a skewed distribution, with most participants performing at ceiling, we created a weighted score (1-5) based on accuracy as follows: 1 = <35 correct; 2 = 35-41 correct; 3 = 42 correct; 4 = 43 correct; 4.25 = 44 correct; 4.5 = 45 correct; 4.75 = 46 correct; 5 = 47-48 correct. Next, we subtracted SYM from 10 to inverse the variable such that higher scores indicated better performance in order to combine the variable with SLS performance for the composite (in the original variable, faster, i.e., lower, scores indicated better performance). We selected 10 seconds as the time to subtract from because nearly all participants had an average correct items response time of less than 10 seconds when this composite was *a priori* conceptualized (April 30, 2022). Then, we multiplied the rescaled SYM variable by the accuracy-weighted score to compute an accuracy-weighted average correct item response time variable (SYM_aw_); for the few participants for whom SYM was >10 seconds, participants received 0 on the SYM_aw_ variable. Finally, to complete calculation of the MTD-SBCr, we added SLS Sum of Trials and the SYM_aw_ variable. As expected, MTD-SBCz and MTD-SBCr are highly correlated (r=0.98).

### In-Person Cognitive Measures

All participants completed an in-person neuropsychological evaluation that was administered by a psychometrist and supervised by a board-certified neuropsychologist (JAF or MMM). The evaluation includes nine neuropsychological tests that together comprise four cognitive domains. The cognitive domains and their constituent tests are as follows: *Memory*-Auditory Verbal Learning Test (AVLT) delayed recall^33^ and Wechsler Memory Scale-Revised Logical Memory II & Visual Reproduction II;^34^ *Language –* Boston Naming Test^35^ and Category Fluency;^36^ *Executive Function –* Trail Making Test B^37^ and Wechsler Adult Intelligence Scale-Revised (WAIS-R) Digit Symbol Coding subtest;^38^ and *Visuospatial Skills –* WAIS-R Picture Completion and Block Design subtests.^38^ Given interest in comparing MTD cognitive outcomes with comparable in-person neuropsychological measures, we only included performances on the AVLT, Trail Making Test B, and WAIS-R Digit Symbol Coding in our subtest analyses.

Additionally, two in-person composite cognitive measures were calculated from the above neuropsychological test data. First, the Mayo Clinic Preclinical Alzheimer’s Cognitive Composite (Mayo-PACC) is an average of z-scores from the AVLT sum of trials, Trail Making Test B (reversed) and animal fluency.^14^ Second, a Global z-score was calculated as the average of z-scores from all nine neuropsychological tests; this is routinely used in MCSA publications (see Stricker et al.^14^ for comparison to several PACCs). Like for MTD-SBCz, both Mayo-PACC and Global-z composites were calculated using participants from the full sample available at the time of this analyses (participants with and without available neuroimaging) as the reference group.

Finally, the Short Test of Mental Status (STMS) is a brief, multi-domain cognitive screening measure (38 points possible) that was administered to all participants by the study physician.^39^ The STMS has been shown to have greater ability to detect MCI relative to the MMSE^40^ and the MoCA^41^ and is included for reference.

## Neuroimaging Acquisition & Processing

### Amyloid PET

Amyloid PET scans were acquired via a GE Discovery RX or DXT PET/CT scanner using the ^11^C-Pittsburgh Compound B (PiB) ligand as previously described.^42^ A PiB PET standardized uptake value ratio (SUVR) was defined for each participant by computing the median PiB uptake in gray and white matter of the prefrontal, orbitofrontal, parietal, temporal, anterior cingulate, and posterior cingulate/precuneus regions of interest (ROI) and dividing this by the median uptake in the cerebellar crus gray matter. Together, this continuous SUVR metric is referred to as the amyloid PET meta-ROI and was our measure of interest for amyloid burden. This measure was natural log transformed and z-scored for analyses to facilitate comparisons of results.

### Tau PET

Tau PET scans were also acquired using a GE Discovery RX or DXT PET/CT scanner using the flortaucipir (^18^F-AV-1451) ligand as previously described.^43,44^ Two tau PET metrics were used for the purposes of this study. First, flortaucipir PET SUVR was calculated using the median uptake in gray and white matter of the entorhinal, amygdala, parahippocampal, fusiform, inferior temporal, and middle temporal ROIs divided by the median uptake in the cerebellar crus gray matter; this metric is referred to as the tau PET meta-ROI as a more global measure of tau burden. Second, given previous associations between flortaucipir uptake in the entorhinal cortex (EC) and PiB in CU participants in our sample,^45^ we also investigated this region individually, which is referred to as Tau PET EC-ROI. Both SUVR metrics were investigated continuously and were natural log transformed and z-scored for analysis.

### Hippocampal Volume

MRI scans were conducted on a Siemens 3T Prisma scanner using a 3D Magnetization Prepared Rapid Acquisition Gradient-Echo (MPRAGE) sequence. As previously described,^10,46^ SPM12 Unified Segmentation was used for tissue-class segmentation with Mayo Clinic Adult Lifespan Template (MCALT; https://www.nitrc.org/projects/mcalt/) settings optimized for the study population. Advanced Normalization Tools (ANTs) symmetric normalization was used to warp the MCALT-ADIR122 atlas for computing intracranial volume (ICV) as well as HV.^47,48^ To adjust HV for ICV, residual values from the linear regression of ICV (x) and HV (y) were calculated with sex-specific formulas as previously described.^10^ Values were additionally natural log transformed and z-scored.

### White Matter Hyperintensities

For quantifying WMH, 3D T2-weighted fluid attenuated inversion recovery (FLAIR) MRI sequences were acquired and co-registered to MPRAGE images.^49,50^ WMH was divided by ICV and multiplied to create a percentage WMH volume, which was natural log transformed due to data skewness and then z-scored.

## Statistical Analyses

Analyses were conducted in SAS version 9.4 (SAS Institute Inc., Cary, NC), with a two-tailed p-value of ≤.05 for significance. Demographics and other participant characteristics for the full sample and by diagnostic categories were summarized using means and standard deviations for continuous data and by counts and percentages for categorical data. Basic between-group differences were calculated using independent-samples t-test for continuous variables and χ^2^ or Fisher tests for categorical variables. Linear regression was used to additionally adjust for age, sex, and education for between-group differences by clinical diagnosis for cognitive data. Hedge’s g was used to calculate effect sizes for between-group differences of continuous data.

For primary analyses, multivariable linear regression was used to investigate associations between five neuroimaging biomarkers, including amyloid PET meta-ROI, tau PET meta-ROI, tau PET EC-ROI SUVRs, HV, and percentage WMH volume (analyzed separately), and remote (MTD-SBCr and MTD-SBCz) and in-person (Mayo-PACC, Global z, STMS) composite indices of cognition while adjusting for age, sex, and education. In order to visually compare the mean estimates from each linear regression model, we opted to z-score these outcomes, with the full sample as the referent group. Multivariable linear regression was also used to analyze associations between the five neuroimaging biomarkers (analyzed separately) and remote and in-person cognitive measures of memory (SLS Sum of Trials, Maximum Learning Span, Trial 1-5 Total, and Short Delay Total and AVLT Sum of Trials) and processing speed/executive function (SYM, SYM_aw_, Trail Making Test B, and Digit Symbol Coding).

Finally, Spearman correlation coefficients were utilized to examine unadjusted associations between imaging biomarkers and remote and in-person cognitive measures, both within the full sample and within diagnostic category.

## Results

### Participant Characteristics

Based on study inclusion criteria, 684 individuals comprised the study sample. The average age at completion of MTD was 70.36 years (11.20 SD), and average time from neuroimaging visit to completion of MTD was 5.75 months (11.39 SD). Approximately 50.3% of the sample was female and participants had about 16 years of education on average. Of study participants, 643 were CU (94%), 34 were diagnosed with MCI (5%); and 7 were diagnosed with dementia (1%) based on consensus conference. Given the small number of participants with dementia, MCI and dementia diagnostic groups were combined in *Table 1*. There were differences between diagnostic groups in terms of age (p<.001) and education (p=.002). Nearly all participants completed MTD remotely (99.6%), with a small number of participants electing to complete MTD in clinic (n=3). For additional participant details, see *Table 1*.

Additionally, Spearman correlation coefficients between remote and in-person cognitive tests show significant associations (all *p*’s < .001; see *Table S1* for full details). Primary associations of interest were robust including associations between MTD-SBCr and other global measures of cognition (Mayo-PACC r=0.68; Global-z r=0.67; Kokmen STMS r=0.56), the SLS and AVLT Sum of Trials (r=0.61), and SYM_aw_ and other measures of processing speed/executive function (Trails B r=-.61; Digit Symbol Coding r=0.59).

## Associations of Neuroimaging Biomarkers and Remote and In-Person Cognitive Measures

### Composite Cognitive Measures

In models adjusted for age, sex, and education, all neuroimaging biomarkers (amyloid meta-ROI, tau meta-ROI, tau EC-ROI, HV, and WMH volume) were significantly associated with the remote measures including the MTD-SBCr and MTD-SBCz (all p’s<.001). When investigating similar associations with in-person cognitive measures, the amyloid meta-ROI (p’s≤.03), tau EC-ROI (p’s≤.03), HV (p’s<.001), and WMH volume (p’s<.001), but not tau meta-ROI (p’s≥.10), were associated with both Mayo-PACC and Global Cognition Z. All neuroimaging biomarkers were significantly associated with the STMS (p’s≤.03). For additional details, see *Table 2*.

**Table 2.** Association Between Standardized Neuroimaging Biomarkers and Composite Cognitive Outcomes Adjusting for Age, Sex, and Education using Linear Regression Models.

| **Cognitive Measure** | **Amyloid PET Meta-ROI SUVR** |  | **Tau PET Meta-ROI SUVR** |  | **Tau PET EC-ROI SUVR** |  | **Hippocampal Volume z-score** |  | **% WMH Volume, ln** |  |
| --- | --- | --- | --- | --- | --- | --- | --- | --- | --- | --- |
|  | Mean estimate (95% CI) | p | Mean estimate (95% CI) | p | Mean estimate (95% CI) | p | Mean estimate (95% CI) | p | Mean estimate (95% CI) | p |
| ***Remote*** |  |  |  |  |  |  |  |  |  |  |
| MTD-SBCr | -0.15<br>(-0.23, -0.08) | <.001 | -0.19<br>(-0.25, -0.13) | <.001 | -0.24<br>(-0.30, -0.17) | <.001 | 0.24<br>(0.17, 0.31) | <.001 | -0.19<br>(-0.28, -0.09) | <.001 |
| MTD-SBCz | -0.16<br>(-0.24, -0.09) | <.001 | -0.20<br>(-0.26, -0.13) | <.001 | -0.24<br>(-0.31, -0.18) | <.001 | 0.25<br>(0.18, 0.32) | <.001 | -0.19<br>(-0.28, -0.09) | <.001 |
| ***In-Person*** |  |  |  |  |  |  |  |  |  |  |
| Mayo-PACC | -0.14<br>(-0.21, -0.06) | <.001 | -0.05<br>(-0.12, 0.01) | 0.10 | -0.10<br>(-0.16, -0.03) | 0.005 | 0.20<br>(0.13, 0.27) | <.001 | -0.17<br>(-0.26, -0.08) | <.001 |
| Global Cognition Z | -0.09<br>(-0.16, -0.01) | 0.03 | -0.00<br>(-0.07, 0.07) | 0.99 | -0.08<br>(-0.15, -0.01) | 0.03 | 0.21<br>(0.14, 0.28) | <.001 | -0.15<br>(-0.24, -0.06) | <.001 |
| STMS | -0.11<br>(-0.21, -0.01) | 0.03 | -0.23<br>(-0.31, -0.15) | <.001 | -0.23<br>(-0.32, -0.14) | <.001 | 0.30<br>(0.21, 0.40) | <.001 | -0.18<br>(-0.30, -0.06) | 0.004 |

Note: All cognitive measures and imaging biomarkers were additionally z-scored to facilitate estimate comparisons within the table. CI = confidence interval; EC = entorhinal cortex; Global Cognition z = average z across all neuropsychological tests administered during an in-person visit; Mayo-PACC = Mayo Preclinical Alzheimer's disease Cognitive Composite (average z of Auditory Verbal Learning Test sum of trials, animal fluency, and Trails B); MTD = Mayo Test Drive; MTD-SBCr = Mayo Test Drive Screening Battery Composite raw, where Stricker Learning Span sum of trials + Symbols Test accuracy-weighted average correct item response time; MTD-SBCz = Mayo Test Drive Screening Battery Composite z; p = p-value; PET = Positron Emission Tomography; ROI = region of interest; STMS = Short Test of Mental Status, which is similar to the MMSE, is included for reference; SUVR = Standard Uptake Volume Ratio; WMH = white matter hyperintensities.

### Individual Cognitive Measures

In investigating the relationship between neuroimaging biomarkers and memory measures, all neuroimaging biomarkers were associated with the remote measure, SLS Sum of Trials (p’s≤.002). All biomarkers were also associated with the in-person measure, AVLT Sum of Trials (p’s≤.004).

For measures of processing speed/executive function, all neuroimaging biomarkers were significantly associated with both SYM and SYM_aw_ (p’s≤.02). For in-person measures, all neuroimaging biomarkers were associated with Trail Making Test B performance (p’s≤.003). Only HV and WMH volume were associated with Digit Symbol Coding (p’s≤.009). For additional details, see *Table 3*.

**Table 3.**
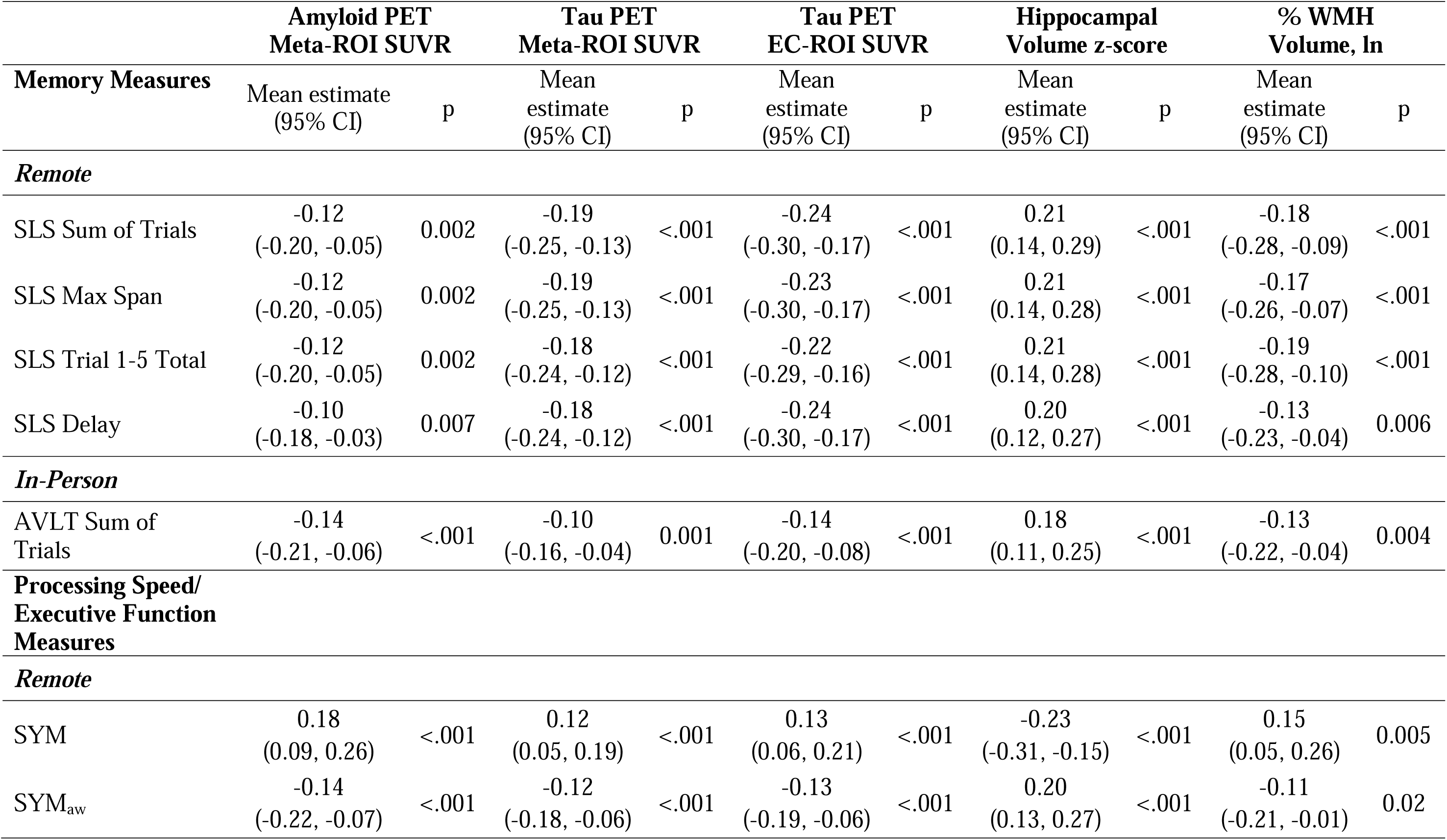
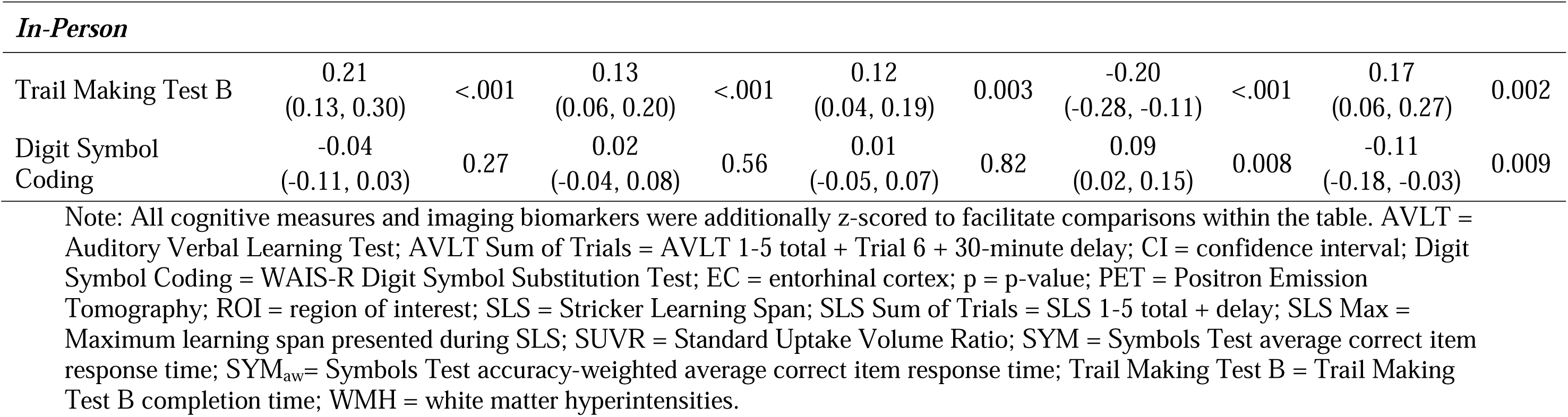
Association Between Standardized Neuroimaging Biomarkers and Cognitive Subtest Outcomes Adjusting for Age, Sex, and Education using Linear Regression Models.

| Memory Measures | Amyloid PET<br>Meta-ROI SUVR |  | Tau PET<br>Meta-ROI SUVR |  | Tau PET<br>EC-ROI SUVR |  | Hippocampal<br>Volume z-score |  | % WMH<br>Volume, ln |  |
| --- | --- | --- | --- | --- | --- | --- | --- | --- | --- | --- |
|  | Mean estimate<br>(95% CI) | p | Mean<br>estimate<br>(95% CI) | p | Mean<br>estimate<br>(95% CI) | p | Mean<br>estimate<br>(95% CI) | p | Mean<br>estimate<br>(95% CI) | p |
| *Remote* |  |  |  |  |  |  |  |  |  |  |
| SLS Sum of Trials | -0.12<br>(-0.20, -0.05) | 0.002 | -0.19<br>(-0.25, -0.13) | <.001 | -0.24<br>(-0.30, -0.17) | <.001 | 0.21<br>(0.14, 0.29) | <.001 | -0.18<br>(-0.28, -0.09) | <.001 |
| SLS Max Span | -0.12<br>(-0.20, -0.05) | 0.002 | -0.19<br>(-0.25, -0.13) | <.001 | -0.23<br>(-0.30, -0.17) | <.001 | 0.21<br>(0.14, 0.28) | <.001 | -0.17<br>(-0.26, -0.07) | <.001 |
| SLS Trial 1-5 Total | -0.12<br>(-0.20, -0.05) | 0.002 | -0.18<br>(-0.24, -0.12) | <.001 | -0.22<br>(-0.29, -0.16) | <.001 | 0.21<br>(0.14, 0.28) | <.001 | -0.19<br>(-0.28, -0.10) | <.001 |
| SLS Delay | -0.10<br>(-0.18, -0.03) | 0.007 | -0.18<br>(-0.24, -0.12) | <.001 | -0.24<br>(-0.30, -0.17) | <.001 | 0.20<br>(0.12, 0.27) | <.001 | -0.13<br>(-0.23, -0.04) | 0.006 |
| *In-Person* |  |  |  |  |  |  |  |  |  |  |
| AVLT Sum of Trials | -0.14<br>(-0.21, -0.06) | <.001 | -0.10<br>(-0.16, -0.04) | 0.001 | -0.14<br>(-0.20, -0.08) | <.001 | 0.18<br>(0.11, 0.25) | <.001 | -0.13<br>(-0.22, -0.04) | 0.004 |
| **Processing Speed/**<br>**Executive Function**<br>**Measures** |  |  |  |  |  |  |  |  |  |  |
| *Remote* |  |  |  |  |  |  |  |  |  |  |
| SYM | 0.18<br>(0.09, 0.26) | <.001 | 0.12<br>(0.05, 0.19) | <.001 | 0.13<br>(0.06, 0.21) | <.001 | -0.23<br>(-0.31, -0.15) | <.001 | 0.15<br>(0.05, 0.26) | 0.005 |
| SYM<sub>aw</sub> | -0.14<br>(-0.22, -0.07) | <.001 | -0.12<br>(-0.18, -0.06) | <.001 | -0.13<br>(-0.19, -0.06) | <.001 | 0.20<br>(0.13, 0.27) | <.001 | -0.11<br>(-0.21, -0.01) | 0.02 |
| ***In-Person*** |  |  |  |  |  |  |  |  |  |  |
| Trail Making Test B | 0.21<br>(0.13, 0.30) | <.001 | 0.13<br>(0.06, 0.20) | <.001 | 0.12<br>(0.04, 0.19) | 0.003 | -0.20<br>(-0.28, -0.11) | <.001 | 0.17<br>(0.06, 0.27) | 0.002 |
| Digit Symbol<br>Coding | -0.04<br>(-0.11, 0.03) | 0.27 | 0.02<br>(-0.04, 0.08) | 0.56 | 0.01<br>(-0.05, 0.07) | 0.82 | 0.09<br>(0.02, 0.15) | 0.008 | -0.11<br>(-0.18, -0.03) | 0.009 |

Note: All cognitive measures and imaging biomarkers were additionally z-scored to facilitate comparisons within the table. AVLT = Auditory Verbal Learning Test; AVLT Sum of Trials = AVLT 1-5 total + Trial 6 + 30-minute delay; CI = confidence interval; Digit Symbol Coding = WAIS-R Digit Symbol Substitution Test; EC = entorhinal cortex; p = p-value; PET = Positron Emission Tomography; ROI = region of interest; SLS = Stricker Learning Span; SLS Sum of Trials = SLS 1-5 total + delay; SLS Max = Maximum learning span presented during SLS; SUVR = Standard Uptake Volume Ratio; SYM = Symbols Test average correct item response time; SYM<sub>aw</sub> = Symbols Test accuracy-weighted average correct item response time; Trail Making Test B = Trail Making Test B completion time; WMH = white matter hyperintensities.

### Comparing MTD Performances Between Diagnostic Groups

As anticipated, differences were observed in MTD performance by diagnostic category. Specifically, participants in the MCI/dementia category demonstrated worse performances across all MTD measures relative to participants in the CU category (all p’s<.001), with large effect sizes for both MTD composites (Hedges g=-2.02 and –2.10), the memory measure, SLS Sum of Trials (Hedges g=-1.78), and processing speed/executive function measures including both Symbols measures (Hedges g=-1.65 and 1.91). Group differences remained significant after adjusting for age, sex, and education (all p’s≤.003; see *Table 1*). Supplementary analyses (see *Table S2*) show that these findings are similar when limited to participants in the CU versus MCI categories, with all *p*’s<.001 and large effect sizes (Hedges g=-1.83 and –1.90) for MTD composite measures, SLS Sum of Trials (Hedges g=-1.62), and for both variants of the Symbols measure (Hedges g=-1.46 and 1.69). Although results must be viewed cautiously given the small number of dementia participants (N=7), preliminary findings comparing MTD performances between those diagnosed with MCI versus dementia suggest a further decrease in performance with increased disease severity. Those diagnosed with dementia showed lower MTD performances for the MTD composite measures and SLS Sum of Trials (*p*’s≤.03; Hedges g=-0.94 to –1.08); Symbols measures approached significance (p’s=.06; Hedges g =-0.81 to 0.82). See *Table S2* for additional details.

### Correlations between Imaging Biomarkers and Remote and In-Person Cognitive Tests by Diagnostic Category

Examination of Spearman correlation coefficients between imaging biomarkers and cognitive measures revealed that for individuals who were diagnosed with either MCI or dementia, correlations were generally stronger when compared with CU participants for both remote and in-person cognitive measures. Please see *Table 4* and *Figure 1* for additional details. For correlations between imaging biomarkers and cognitive measures in the full sample, please see *Table S3*.

**Figure 1.**
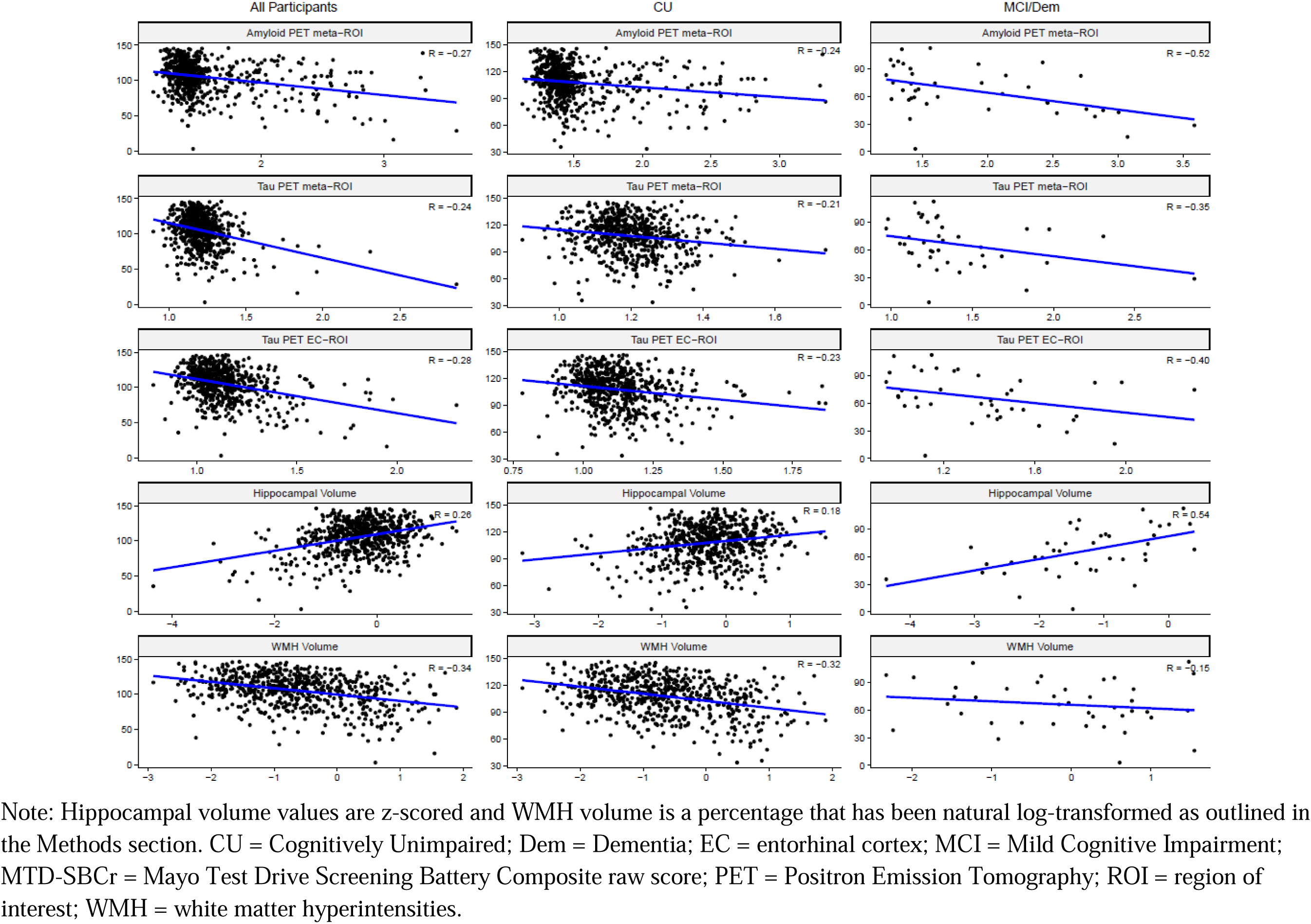
Scatterplots depicting associations between the MTD raw screening battery composite (MTD-SBCr) score (y-axis) and each imaging variable (x-axis) for the full sample (left panel), in cognitively unimpaired participants (center panel), and in participants with MCI or dementia (right panel).

**Table 4.**
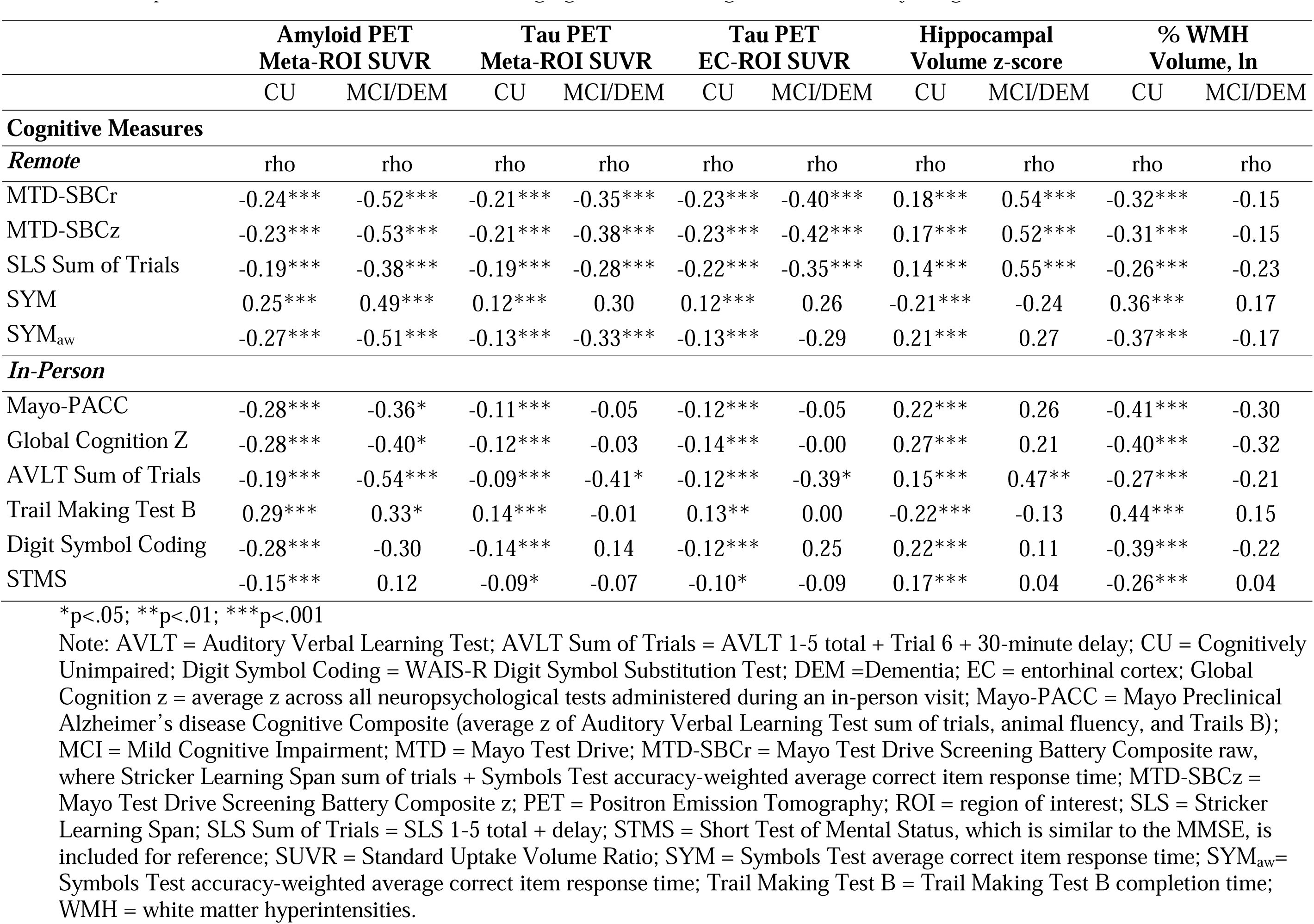
Spearman Correlations between Neuroimaging Metrics and Cognitive Measures by Diagnosis.

## Discussion

In this study of nearly 700 older adults who were predominantly CU, we showed that worse performances on MTD, as measured via a raw composite score (MTD-SBCr) derived from the SLS and Symbols tests, was significantly associated with higher levels of amyloid in a PET meta-ROI, higher levels of tau in the entorhinal cortex specifically as well as in a PET meta-ROI, lower HV, and higher WMH. There were also significant associations across all imaging biomarkers with MTD-SBCz, as well as the SLS Sum of Trials, SYM, and SYM_aw_ metrics. For reference, Mayo-PACC and Global Cognition z, two person-administered cognitive composites, were also associated with all imaging biomarkers with the exception of the tau PET meta-ROI. The AVLT (similar to the SLS) and Trail Making Test B (similar to the Symbols test) were both significantly associated with all imaging biomarkers, whereas Digit Symbol Coding (also similar to the Symbols test) was only associated with HV and WMH. Additionally, we demonstrated that MTD performances for both composites (MTD-SBCr and MTD-SBCz) differ by diagnostic category (CU, MCI, dementia) in an expected fashion, and that associations between MTD and imaging biomarkers are generally stronger for those diagnosed with MCI or dementia compared with CU individuals, again as expected. MTD and in-person neuropsychological measures were robustly associated with each other, as well. Together, these findings highlight MTD’s ability to detect cognitive change in a large group of predominantly (94%) CU individuals that is associated with greater levels of amyloid and tau burden, consistent with the biological diagnosis of AD, as well as hippocampal and WMH volumes, together which supports MTD’s criterion validity. Given that the MTD-SBCr additionally demonstrated the ability to differentiate diagnostic groups, it has promising utility as a generalizable tool for detecting subtle objective cognitive impairment that is independent of study-specific samples. Overall, findings from this study highlight the ability of MTD to detect subtle cognitive change associated with biomarkers of AD and neurodegeneration across diagnostic groups in a completely remote, self-administered digital fashion.

Remote, digital cognitive assessments have been developing more rapidly over the past few years as older adults have become more accustomed to digital technology, as its effectiveness has been shown in reaching large groups of people who may otherwise have limited access to cognitive assessment, and due to its necessity during the COVID-19 pandemic.^31^ To date, most of these remote digital assessments rely on multiple assessments, either once per day over several days (e.g., Boston Remote Assessment for Neurocognitive Health; BRANCH),^51^ or multiple times per day over several days (e.g., Mobile Monitoring of Cognitive Change; M2C2).^52^ Performance on BRANCH has robust associations with the PACC5 as well as associations with cortical amyloid and entorhinal tau deposition,^51^ with diminished seven-day practice effects on BRANCH linked to increased levels of amyloid and declines on the PACC5 over one year in CU individuals.^53^ Similarly, diminished practice effects on C3 over a three month period have been associated with greater amyloid and tau burden as well as steeper annual decline on the PACC5.^54^ Another remote digital assessment – the M2C2 Prices task as part of the NIH Mobile Toolbox – distinguishes A+ versus A-participants better than the MoCA.^52^ Our group has previously shown that a one-time administered SLS discriminates between A+ and A-groups, as well as A+T+ and A-T-groups in a predominantly CU sample.^11^

In this study, we further demonstrated that worse performance on the MTD-SBCr and MTD-SBCz were both continuously associated with higher amyloid, tau, and WMH burden, and lower HV. By including these additional biomarkers in this study of predominantly CU individuals, we were able to demonstrate the sensitivity of MTD in detecting subtle cognitive change was not only associated with amyloid, but with other AD-related biomarkers, as well. Importantly, MTD only requires a one-time administration, rather than repeated measurements over days to months, as other remote digital assessments require currently, and is still able to detect these biomarker-associated cognitive changes in CU individuals. Additionally, MTD’s associations with these imaging biomarkers are broadly comparable to the continuous associations between the in person-administered Mayo-PACC and imaging biomarkers.

The components that comprise the two MTD composites – the SLS and Symbols Test – were both individually associated with all imaging biomarkers investigated in this study. These associations were similar to their person-administered cognitive test counterparts that were provided for reference in this study – the AVLT, Trail Making Test B, and Digit Symbol Coding, with the AVLT and Trail Making Test B also associated with all imaging biomarkers, and Digit Symbol Coding associated with HV and WMH. We did not directly compare associations between MTD and imaging biomarkers with traditional person-administered cognitive measures and imaging biomarkers, as the SLS has already been directly compared to the AVLT in its ability to predict dichotomous AD biomarker status.^11^ Our goal was to investigate continuous associations, for which we prioritized inclusion of all participants with imaging data as opposed to limiting the sample to those with all cognitive measures which would have excluded most individuals with dementia due to test battery differences across the MCSA and ADRC. In particular, most dementia participants are missing data for Global Cognition z given differences in MCSA and ADRC test batteries.

One challenge with PACCs and current remote digital cognitive assessments is that findings are typically study-specific, with results typically depicted as z-scores derived from the sample itself. We developed the MTD-SBCr to move beyond study-specific scores and cutoffs to develop a score that can be interpreted independent of study sample. We demonstrated that the MTD-SBCr, which does not rely on study-specific z-scores, was able to distinguish between CU, MCI, and dementia diagnostic groups with large effect sizes. Additionally, when separated by diagnostic groups, MTD was significantly correlated with all imaging biomarkers, with stronger associations for MCI and dementia diagnostic categories. As such, the MTD-SBCr is sensitive to diagnostic categories associated with AD-related biomarkers. Given that it does not rely on study-specific performances like z-scores for interpretation, the MTD-SBCr could be used similarly to an MMSE or MoCA, where the raw score in and of itself has meaning associated with broad diagnostic categories (i.e., cognitively impaired versus unimpaired). This index could also be useful because it does not adjust for age-related effects. We now know that age-related cognitive change may actually be due to previously undetected neuropathological change^17,55^, so adjusting for age-related associations may subtract meaning from cognitive scores. As such, the MTD-SBCr would allow for understanding of cognitive status in clinical settings independent of these factors, and could allow for early screening, triage, and routing for specialty care. Future work will develop clinical cut-offs based on sensitivity and specificity for MTD-SBCr.

There are several limitations to this study. Importantly, the MCSA is a population-based study that represents the demographic makeup of Rochester, MN, which is primarily comprised of non-Hispanic white individuals. As such, inclusion of individuals from underrepresented racial and ethnic groups is a clear weakness. We are actively expanding the use of MTD to underrepresented groups and developing a Spanish adaptation of MTD to address this important limitation. Additionally, while the focus of this study was detecting biomarker-cognition associations in the earlier clinical stages of the AD continuum, inclusion of more participants with MCI and/or dementia could have allowed for even stronger associations between AD-related biomarkers and MTD performance and may have allowed for better characterization of MTD performance in these diagnostic groups. It is also important to note that the remote nature of MTD allows for variability in the testing environment, possible interference, unknown trouble navigating the technology, or the possibility of unanticipated assistance during the assessment. Future work is needed to examine the potential impact of these factors on results.

This study has notable strengths. We were able to examine associations between multiple imaging biomarkers of Alzheimer’s disease and performance on a remote, digital cognitive assessment composite measure in a relatively large sample. Further, our ability to demonstrate continuous associations between these biomarkers and cognitive performance in a predominantly CU sample suggests promising ability to detect subtle cognitive changes at the earliest stages of the AD biological cascade. Given that MTD is a one-time, approximately 15-minute remote assessment, this provides great utility for access and ease for inclusion in clinical trials as well as in clinical settings more broadly. The development of the MTD-SBCr augments this utility, as it provides greater ease of interpretation for MTD performance in a similar fashion to current paper-and-pencil cognitive screening tools, such that clinicians could easily implement this into their practice in the future. Furthermore, the nature of the MCSA, which is population based, provides generalizability of our findings because individuals with other comorbidities are not excluded. Our findings are also responsive to the update to the amyloid-tau-neurodegeneration (ATN) research framework,^13^ as we demonstrate MTD’s association with core AD biomarkers including amyloid and tau PET, HV – a non-specific neurodegeneration biomarker, and WMH indicating common non-AD vascular co-pathology. To our knowledge, demonstrating associations with all of these biomarkers within a single study has not been completed to date with other remote digital assessments.

In summary, our study demonstrates that the MTD composites, comprised of the SLS and the Symbols Test, are associated with continuous measures of imaging biomarkers including amyloid and tau PET, HV, and WMH. These associations are also seen in the individual subtests of MTD – SLS and Symbols Test. Further, the MTD composite differentiates CU, MCI, and dementia diagnostic groups, and associations between MTD and imaging biomarkers are strengthened in MCI and dementia diagnostic groups. Together, our findings highlight the criterion validity of MTD as a remote, digital cognitive assessment linked with AD-related biological change that can be detected even in individuals who are not demonstrating any overt cognitive impairment.

## Data Availability

All data produced in the present study are available upon reasonable request to the authors.

## Acknowledgements

The authors wish to thank the participants and staff at the Mayo Clinic Study of Aging and Mayo Alzheimer’s Disease Research Center.

