## Supplementary material for "Continuous Associations Between Remote Self-Administered Cognitive Measures and Imaging Biomarkers of Alzheimer’s Disease": Table S1

Supplemental Online Resources

*medRxiv*

Elizabeth A. Boots, Ph.D.,[1] Ryan D. Frank, M.S.,[2] Winnie Z. Fan, M.S., [2] Teresa J. Christianson, M.S., [2] Walter K. Kremers, Ph.D., [2] John L. Stricker, Ph.D., [3] Mary M. Machulda, Ph.D., [1] Julie A. Fields, Ph.D., [1] Jason Hassenstab, PhD., [4] Jonathan Graff-Radford, M.D., [5] Prashanthi Vemuri, Ph.D., [6] Clifford R. Jack, M.D.,[6] David S. Knopman, M.D., [5] Ronald C. Petersen, M.D., Ph.D., [5] Nikki H. Stricker, Ph.D.[1]

[1]Divison of Neurocognitive Disorders, Department of Psychiatry and Psychology, Mayo Clinic, Rochester, MN, USA
[2]Division of Biomedical Statistics and Informatics, Department of Quantitative Health Sciences, Mayo Clinic, Rochester, Minnesota, USA
[3] Department of Information Technology, Mayo Clinic, Rochester, Minnesota, USA
[4]Department of Neurology and Psychological & Brain Sciences, Washington University in St. Louis, St. Louis, Missouri, USA
[5] Department of Neurology, Mayo Clinic, Rochester, Minnesota, USA
[6] Department of Radiology, Mayo Clinic, Rochester, Minnesota, USA

Copyright 2024 Mayo Foundation for Medical Education and Research.

A poster presentation of early/partial data from this work in a smaller sample size was presented at the Alzheimer's Association International Conference (July 2023). Another portion of this work was submitted as an abstract for the American Psychological Association Annual Convention (August 2024).

*Supplementary Table 1*. Spearman Correlations between remote and in-person cognitive measures.

| | Remote Mayo Test Drive Measures | | | | | | | | | |
|---|---|---|---|---|---|---|---|---|---|---|
| | MTD-SBCr | | MTD-SBCz | | SLS Sum of Trials | | SYM | | $SYM_{aw}$ | |
| **In-Person Cognitive Measures** | rho | N | rho | N | rho | N | rho | N | rho | N |
| Mayo-PACC | 0.68 *** | 657 | 0.68 *** | 657 | 0.60 *** | 658 | -0.60 *** | 658 | 0.59 *** | 658 |
| Global Cognition Z | 0.67 *** | 626 | 0.67 *** | 626 | 0.59 *** | 627 | -0.59 *** | 627 | 0.58 *** | 627 |
| AVLT Sum of Trials | 0.64 *** | 668 | 0.65 *** | 668 | 0.61 *** | 669 | -0.42 *** | 669 | 0.43 *** | 669 |
| Trail Making Test B | -0.57 *** | 665 | -0.57 *** | 664 | -0.46 *** | 666 | 0.62 *** | 666 | -0.61 *** | 666 |
| Digit Symbol Coding | 0.55 *** | 654 | 0.56 *** | 654 | 0.44 *** | 655 | -0.63 *** | 655 | 0.59 *** | 655 |
| STMS | 0.56 *** | 673 | 0.55 *** | 673 | 0.49 *** | 674 | -0.49 *** | 674 | 0.50 *** | 674 |

*p<.05; **p<.01; ***p<.001

Note: AVLT = Auditory Verbal Learning Test; AVLT Sum of Trials = AVLT 1-5 total + Trial 6 + 30-minute delay; Digit Symbol Coding = WAIS-R Digit Symbol Substitution Test; Global Cognition z = average z across all neuropsychological tests administered during an in-person visit; Mayo-PACC = Mayo Preclinical Alzheimer's disease Cognitive Composite (average z of Auditory Verbal Learning Test sum of trials, animal fluency, and Trails B); MTD-SBCr = Mayo Test Drive Screening Battery Composite raw, where Stricker Learning Span sum of trials + Symbols Test accuracy-weighted average correct item response time; MTD-SBCz = Mayo Test Drive Screening Battery Composite z; SLS = Stricker Learning Span; SLS Sum of Trials = SLS 1-5 total + delay; STMS = Short Test of Mental Status, which is similar to the MMSE, is included for reference; SYM = Symbols Test average correct item response time; $SYM_{aw}$= Symbols Test accuracy-weighted average correct item response time; Trail Making Test B = Trail Making Test B completion time.

*Supplemental Table 2.* Participant Characteristics with Comparisons between Cognitively Unimpaired, Mild Cognitive Impairment, and Dementia Participants

| **Characteristic*** | **CU** | **MCI** | **CU vs MCI** | | **Dementia** | **CU vs Dementia** | | **MCI vs Dementia** | |
|---|---|---|---|---|---|---|---|---|---|
| | N=643 | N=34 | p | Hedge's g | N=7 | p | Hedge's g | p | Hedge's g |
| **Demographics** | | | | | | | | | |
| Age at MCSA/ADRC visit, years | 69.890 (11.111) | 78.809 (10.256) | <0.001† | 0.81 (0.46, 1.15) | 72.820 (8.148) | 0.49† | 0.26 (-0.48, 1.01) | 0.16† | -0.60 (-1.43, 0.22) |
| Sex, N (%) Female | 318 (49.5%) | 19 (55.9%) | 0.47‡ | -- | 3 (42.9%) | 1.00‡ | -- | 0.68‡ | -- |
| Education, years | 15.719 (2.335) | 14.206 (2.649) | <0.001† | -0.64 (-0.99, -0.30) | 16.000 (3.317) | 0.75† | 0.12 (-0.62, 0.86) | 0.13† | 0.65 (-0.18, 1.48) |
| Race, N (%) White | 624 (97.0%) | 33 (97.1%) | 1.00§ | -- | 7 (100.0%) | 0.64§ | -- | 1.00§ | -- |
| Ethnicity, N (%) Non-Hispanic | 639 (99.4%) | 34 (100.0%) | 1.00§ | -- | 7 (100.0%) | 1.00§ | -- | -- | -- |
| In-person visit to MTD, months | 0.709 (1.896) | 1.308 (3.093) | 0.08† | 0.30 (-0.04, 0.65) | 0.493 (0.404) | 0.76† | -0.11 (-0.86, 0.63) | 0.49† | -0.29 (-1.10, 0.53) |
| MTD to imaging, months | 6.113 (11.622) | -0.447 (2.988) | 0.001† | -0.58 (-0.92, -0.23) | 2.079 (5.102) | 0.36† | -0.35 (-1.09, 0.40) | 0.08† | 0.74 (-0.09, 1.57) |
| MTD completed in clinic, N (%) | 3 (0.5%) | 0 (0.0%) | 1.00§ | -- | 0 (0.0%) | 1.00§ | -- | -- | -- |
| **Cognition: Mayo Test Drive**[1]‖ | | | | | | | | | |
| MTD-SBCr, n=680 | 107.653 (20.171) | 70.730 (21.322) | <0.001† | -1.83 (-2.19, -1.46) | 45.492 (33.013) | <0.001† | -3.06 (-3.82, -2.29) | 0.01† | -1.07 (-1.92, -0.22) |
| MTD-SBCz, n=680 | 0.030 (0.815) | -1.525 (0.922) | <0.001† | -1.90 (-2.26, -1.53) | -2.629 (1.433) | <0.001† | -3.23 (-4.00, -2.47) | 0.01† | -1.08 (-1.93, -0.23) |

| | | | | | | | | | |
|---|---|---|---|---|---|---|---|---|---|
| SLS Sum of Trials, n=681 | 75.833 (17.174) | 48.030 (17.853) | <0.001† | -1.62 (-1.98, -1.26) | 30.857 (20.708) | <0.001† | -2.61 (-3.37, -1.85) | 0.03† | -0.94 (-1.78, -0.09) |
| SYM, n=681 | 3.337 (1.067) | 5.276 (2.207) | <0.001† | 1.69 (1.33, 2.05) | 7.261 (3.358) | <0.001† | 3.53 (2.77, 4.30) | 0.06† | 0.82 (-0.02, 1.65) |
| $SYM_{aw}$, n=681 | 31.839 (6.065) | 22.700 (9.277) | <0.001† | -1.46 (-1.82, -1.10) | 14.635 (13.158) | <0.001† | -2.79 (-3.55, -2.03) | 0.06† | -0.81 (-1.64, 0.03) |
| **Cognition: In-Person Measures**[2‖] | | | | | | | | | |
| Mayo-PACC, n=661 | 0.026 (0.738) | -1.712 (1.009) | <.001† | -2.31 (-2.69, -1.93) | -1.502 (0.779) | <.001† | -2.07 (-3.06, -1.08) | 0.49† | 0.21 (-0.83, 1.25) |
| Global Cognition z, n=629 | 1.518 (0.887) | -0.871 (0.819) | <.001† | -2.70 (-3.12, -2.28) | -- | -- | -- | -- | -- |
| AVLT Sum of Trials, n=668 | 66.979 (17.568) | 36.813 (11.255) | <.001† | -1.74 (-2.11, -1.37) | 27.167 (6.113) | 0.001† | -2.27 (-3.09, -1.46) | 0.81† | -0.90 (-1.80, -0.01) |
| Trail Making Test B, n=669 | 72.472 (34.875) | 158.469 (82.729) | <.001† | 2.24 (1.86, 2.61) | 150.200 (101.751) | <.001† | 2.18 (1.29, 3.07) | 0.49† | -0.10 (-1.04, 0.85) |
| Digit Symbol Coding, n=658 | 52.060 (12.222) | 34.179 (9.843) | <.001† | -1.47 (-1.86, -1.09) | -- | -- | -- | -- | -- |
| STMS, n=677 | 35.962 (1.905) | 30.412 (2.797) | <.001† | -2.83 (-3.21, -2.46) | 24.333 (8.066) | 0.004† | -5.74 (-6.60, -4.87) | 0.02† | -1.55 (-2.49, -0.62) |
| **Neuroimaging Metrics**[1] | | | | | | | | | |
| Amyloid PET Meta-ROI SUVR, n=670 | 1.527 (0.341) | 1.780 (0.570) | 0.09† | 0.71 (0.37, 1.06) | 2.423 (0.721) | <.001† | 2.59 (1.83, 3.35) | 0.10† | 1.08 (0.23, 1.93) |
| Tau PET Meta-ROI SUVR, n=667 | 1.197 (0.098) | 1.288 (0.218) | <.001† | 0.84 (0.50, 1.19) | 1.881 (0.574) | <.001† | 6.08 (5.26, 6.89) | 0.04† | 1.97 (1.04, 2.89) |
| Tau PET EC-ROI SUVR, n=667 | 1.120 (0.130) | 1.296 (0.267) | <.001† | 1.26 (0.91, 1.61) | 1.672 (0.403) | <.001† | 4.09 (3.31, 4.87) | 0.09† | 1.29 (0.42, 2.15) |
| Hippocampal Volume z-score, n=680 | -0.283 (0.611) | -1.255 (1.097) | <.001† | -1.51 (-1.86, -1.16) | -1.477 (0.553) | <.001† | -1.96 (-2.71, -1.20) | 0.27† | -0.22 (-1.03, 0.60) |

| | | | | | | | | | |
|---|---|---|---|---|---|---|---|---|---|
| % WMH Volume, ln, n=665 | -0.680 (0.884) | -0.094 (1.060) | 0.41† | 0.66 (0.31, 1.00) | -0.193 (1.058) | 0.21† | 0.55 (-0.20, 1.29) | 0.64† | -0.09 (-0.91, 0.72) |

*Values are presented as Mean (Standard Deviation) unless otherwise noted.
† T-test
‡ Chi-square
§ Fisher
‖ P-values are also <0.001 when adjusted for age, sex, and education.
[1] Mayo Test Drive and Neuroimaging are independent of diagnosis (data not considered for consensus diagnosis).
[2] Results of in-person cognitive measures are considered for consensus diagnosis.
Note: ADRC = Alzheimer's Disease Research Center; AVLT = Auditory Verbal Learning Test; CU = Cognitively Unimpaired; EC = entorhinal cortex; Mayo-PACC = Mayo Preclinical Alzheimer's Disease Cognitive Composite; MCI = Mild Cognitive Impairment; MCSA=Mayo Clinic Study of Aging; MRI = Magnetic Resonance Imaging; MTD = Mayo Test Drive; MTD-SBCr = Mayo Test Drive Screening Battery Composite raw; MTD-SBCz = Mayo Test Drive Screening Battery Composite z; p = p-value; PET = Positron Emission Tomography; ROI = Region of Interest; SLS = Stricker Learning Span; STMS = Short Test of Mental Status; SUVR = Standard Uptake Volume Ratio; SYM = Symbols Test average correct item response time; $SYM_{aw}$= Symbols Test accuracy-weighted average correct item response time; WMH = White Matter Hyperintensities.

*Supplemental Table 3.* Spearman Correlations between Neuroimaging Metrics and Cognitive Measures in the Full Sample

| | **Amyloid PET Meta-ROI SUVR** | | **Tau PET Meta-ROI SUVR** | | **Tau PET EC-ROI SUVR** | | **Hippocampal Volume z-score** | | **% WMH Volume, ln** | |
|---|---|---|---|---|---|---|---|---|---|---|
| **Cognitive Measures** | rho | N | rho | N | rho | N | rho | N | rho | N |
| ***Remote*** | | | | | | | | | | |
| MTD-SBCr | -0.27*** | 666 | -0.24*** | 663 | -0.28*** | 663 | 0.26*** | 676 | -0.34*** | 661 |
| MTD-SBCz | -0.27*** | 666 | -0.24*** | 663 | -0.28*** | 663 | 0.25*** | 676 | -0.33*** | 661 |
| SLS Sum of Trials | -0.22*** | 667 | -0.23*** | 664 | -0.27*** | 664 | 0.22*** | 677 | -0.28*** | 662 |
| SYM | 0.28*** | 667 | 0.16 *** | 664 | 0.17*** | 664 | -0.26*** | 677 | 0.38*** | 662 |
| $SYM_{aw}$ | -0.30*** | 667 | -0.17*** | 664 | -0.18*** | 664 | 0.26 *** | 677 | -0.38*** | 662 |
| ***In-Person*** | | | | | | | | | | |
| Mayo-PACC | -0.30*** | 649 | -0.14*** | 646 | -0.16*** | 646 | 0.28*** | 657 | -0.41*** | 642 |
| Global Cognition Z | -0.29*** | 617 | -0.14*** | 614 | -0.17*** | 614 | 0.31*** | 625 | -0.41*** | 611 |
| AVLT Sum of Trials | -0.22*** | 656 | -0.13*** | 653 | -0.17*** | 653 | 0.21*** | 664 | -0.29*** | 649 |
| Trail Making Test B | 0.31*** | 657 | 0.17*** | 654 | 0.17*** | 654 | -0.27*** | 665 | 0.44*** | 650 |
| Digit Symbol Coding | -0.29*** | 646 | -0.15*** | 643 | -0.13*** | 643 | 0.26*** | 654 | -0.40*** | 639 |
| STMS | -0.18*** | 664 | -0.13*** | 661 | -0.16*** | 661 | 0.24*** | 673 | -0.28*** | 658 |

*p<.05; **p<.01; ***p<.001

Note: AVLT = Auditory Verbal Learning Test; AVLT Sum of Trials = AVLT 1-5 total + Trial 6 + 30-minute delay; CU = Cognitively Unimpaired; Digit Symbol Coding = WAIS-R Digit Symbol Substitution Test; DM =Dementia; EC = entorhinal cortex; Global Cognition z = average z across all neuropsychological tests administered during an in-person visit; Mayo-PACC = Mayo Preclinical Alzheimer's disease Cognitive Composite (average z of Auditory Verbal Learning Test sum of trials, animal fluency, and Trails B); MCI = Mild Cognitive Impairment; MTD-SBCr = Mayo Test Drive Screening Battery Composite raw, where Stricker Learning Span sum of trials + Symbols Test accuracy-weighted average correct item response time; MTD-SBCz = Mayo Test Drive Screening Battery Composite z; PET = Positron Emission Tomography; ROI = region of interest; SLS = Stricker Learning Span; SLS Sum of Trials = SLS 1-5 total + delay; STMS = Short Test of Mental Status, which is similar to the MMSE, is included for reference; SUVR = Standard Uptake Volume Ratio; SYM = Symbols Test average correct item response time; $SYM_{aw}$= Symbols Test accuracy-weighted average correct item response time; Trail Making Test B = Trail Making Test B completion time; WMH = white matter hyperintensities.
